# Neonatal unit human resources: coverage for six cadres and trends for staff-to-baby ratios in 65 neonatal units implementing with NEST360 in Kenya, Malawi, Nigeria, Tanzania

**DOI:** 10.1101/2025.03.24.25324517

**Authors:** Rebecca E. Penzias, Eric O. Ohuma, Opeyemi Odedere, Olabisi Dosunmu, George Okello, Hannah Mwaniki, Robert Tillya, Josephine Shabani, Samuel K. Ngwala, Evelyn Zimba, Morris Ondieki Ogero, Christine Bohne, Olukemi Tongo, Veronica Chinyere Ezeaka, Vincent O. Ochieng, Ekran Rashid, William M. Macharia, John Wainaina, Irabi Kassim, Donat Shamba, Nahya Salim, Grace T. Soko, Msandeni Chiume, Alice Tarus, Edith Gicheha, Julius Thomas, Georgia Jenkins, James H. Cross, Rosemary Kamuyu, Junwei Chen, Simon Cousens, Elizabeth M. Molyneux, Maria Oden, Rebecca Richards-Kortum, Joy E. Lawn, David Gathara, with the Data Collection Learning Collaborative Group

**Author notes:** Corresponding Author: Rebecca E. Penzias, Centre for Maternal, Adolescent, Reproductive, & Child Health, London School of Hygiene & Tropical Medicine, London, UK. Joint senior (Joy E. Lawn, David Gathara).

## Abstract

**Background:** Implementing small and sick newborn care (SSNC) requires skilled health workers; however, there is a shortage, adversely impacting patient outcomes and health worker well-being. There are limited data and no current WHO standards for staff-to-baby ratios in neonatal units in low- and middle-income countries (LMICs) to inform policy, planning, and investment.

**Methods:** In 65 neonatal units (36 in Malawi, 13 in Kenya, 7 in Tanzania, and 9 in Nigeria), a health facility assessment (HFA) for SSNC and government-led quality improvement (QI) processes were implemented. Staffing data were collated from baseline HFA (Sept 2019-March 2021) and mid-2023 HFAs, and quarterly QI processes. The unit of analysis was the neonatal unit with day and night staff-to-baby ratios calculated. Ratios were aggregated overall, by country, by hospital level, and neonatal unit occupancy rates. Staff coverage and skill-mix were also analysed for nurses, doctors, clinical officers, laboratory technicians, data clerks, biomedical technicians, and engineers.

**Results:** For 65 neonatal units, median time between baseline and 2023 HFAs was 31 months (Interquartile Range (IQR) 29-34 months). In 2023, only 3 (5%) neonatal units had zero neonatal ward-specific nurses compared to 8 (12%) at baseline during the day. Between baseline and 2023 HFAs, median nurse-to-baby ratios were 1:6 (IQR 1:3-1:11) during the day and 1:10 (IQR 1:6-1:17) at night, with consistency over time. At baseline, only one third of neonatal units had a doctor providing care, or on-call coverage, at all times of day and night (n=20, 31%), and half of hospitals lacked 24-hour laboratory coverage (n=25, 45%) with no change over time. There were improvements in neonatal data clerk (n=32, 49% to n=58, 89%) and biomedical technician (n=45, 69% to n=56, 86%) coverage between baseline and 2023 HFAs.

**Discussion:** Evaluation revealed variability by country and hospital level, and important shortfalls remain in numbers of staff providing care. Neonatal survival in hospitals requires better staff-to-baby ratios, and more skilled staff. To meet the projected shortfall in the health workforce, governments must invest in training the next generation of health workers.

**WHAT IS ALREADY KNOWN ON THIS TOPIC:**

- Sick neonates can die in minutes and having sufficient skilled staff and specialised technologies is crucial. In low- and middle-income countries (LMICs), there is an acute shortage of health workers with the worst shortages in the poorest countries in Africa and Southern Asia.
- There is growing evidence on the impact of low clinical staffing in hospitals on patient safety, outcomes including mortality and quality of care, and health care acquired infections, although most of this literature is from high-income countries (HIC).
- There are few data to inform international standards for staff-to-baby ratios in LMICs. The World Health Organization (WHO) have not yet published standard ratios. National level standards are available for some countries; in India, South Africa, and UK, nurse-to-baby ratios by level of care vary from 1:2 babies in neonatal intensive care to 1:6 babies in special newborn care units.

**WHAT THIS STUDY ADDS:**

- This study analyses trends in staff-to-baby ratios for nurses and doctors from a large dataset involving 65 neonatal units in four countries (Kenya, Malawi, Nigeria, Tanzania). We found that median nurse-to-baby ratios were lower than national standards available for other settings, especially at night (Day: Median 1:6, IQR 1:3-1:11; Night: Median 1:10, IQR 1:6-1:17), with secondary and tertiary hospitals having even lower nurse-to-baby ratios during the day (Secondary/tertiary: Median 1:9, IQR 1:4-1:14; Primary: Median 1:4, IQR 1:3-1:6).
- This study also examines staff coverage and skill-mix for cadres including both clinical (nurses, doctors, clinical officers) and non-clinical staff (biomedical technicians, engineers, data clerks, and laboratory technicians).

- Clinical staff: Despite improvements over time, some neonatal units (n=3, 5%) still did not have any neonatal ward-specific nurses providing care by the 2023 HFA. In addition, many neonatal units lacked doctors providing care, or on-call coverage, 24-hours per day (n=20, 31%) at baseline, and only half of neonatal units had any paediatricians or neonatologists (n=32, 49%) providing care or supervision with no improvements over time.
- Non-clinical staff: Half of hospitals lacked 24-hour laboratory coverage at baseline (n=25, 45%) with no improvements over time. However, there were improvements in neonatal data clerk (n=32, 49% to n=58, 89%) and biomedical technician (n=45, 69% to n=56, 86%) coverage between baseline and 2023 HFAs.

**HOW THIS STUDY MIGHT AFFECT RESEARCH, PRACTICE OR POLICY:**

- Increased investment is needed to enable staffing levels sufficient for high-quality neonatal care.
- Staff coverage and skill-mix data for all cadres is also foundational to inform health workforce allocation, training, planning, and forecasting.
- More routine data on staff-to-baby ratios are crucial for informing increased resource allocation to transform quality of care, and to inform government resource allocation and for developing national and international recommendations for standardised ratios in neonatal units.

## Background

Achieving Sustainable Development Goal 3 (SDG 3) on well-being requires universal health coverage (UHC) and necessitates more human resources for health.^1^ The World Health Organization’s (WHO) Strategy on Human Resources for Health, “Workforce 2030”, emphasised the need to substantially increase the recruitment, development, training, and retention of human resources for health to accelerate progress towards UHC.^2^ In low- and middle-income countries (LMICs), there is an acute shortage of health workers with the worst shortages in the poorest countries in Africa and Southern Asia.^3^ WHO and the World Bank Global Strategy on human resources for health estimated that globally there were 43.5 million health workers in 2013 and that an additional 14.5 million health workers are needed by 2030 using a needs-based “SDG” index model.^3^ In addition, the WHO Standards for Improving the Quality of Care for Small and Sick Newborns (SSNC) in Health Facilities set standards for the detection and management of medical conditions that require specialised devices, consumables, medicines, and a bundle of technologies for small and sick newborns.^4^

The team should include doctors, neonatal nurses, biomedical technicians/engineers, laboratory technicians, effective hospital management, and other paramedical and support staff.^5^ An often neglected issue is the need for data clerks working specifically on the neonatal unit to collect patient-level data to measure and track quality of care. Nurses comprise the largest group of the health workforce, and are critical in multiple roles that span delivery of life-saving interventions, planning, and coordinating care, and management at all levels of the health system. In 2020, WHO estimated a need for an extra 5.9 million nurses to achieve the global agenda by 2030, with 89% of this staffing gap in low and lower-middle incomes countries, primarily in Africa and Southern Asia.^6^

There is growing evidence on the impact of understaffing in hospitals on missed care and patient safety, outcomes including mortality, quality of care, length of stay, and healthcare-associated infections (a proxy for antimicrobial resistance) although most of this literature is from high-income countries (HIC) settings.^7–12^ Similar evidence has been reported for doctors, support staff, and other cadres. Additional implications of workforce shortages include ill health, burnout, and attrition for health workers.^13–15^

Though low staffing numbers have been linked to adverse outcomes, there are currently no evidence-based global standards for staff-to-baby ratios in SSNC units.^5^ However, some countries have national standards for nurse-to-baby ratios based on level of care. For example, in India, recommendations support nurse-to-baby ratios of 1:3 or 1:4 for special newborn care.^4,16^ In South Africa, recommendations support ratios of 1:6 in standard SSNC or Kangaroo Mother Care (KMC) units, 1:2 or 1:3 for high-dependency rooms, and 1:1 or 1:2 for intensive care units.^4,17^ In the United Kingdom, recommendations support 1:1 for intensive care, 1:2 for high-dependency care, and 1:4 for special newborn care.^4,18^

High-quality SSNC in hospitals requires both more numbers and more skills in neonatal care. However, mere quantity of human resources for health is not enough – improving effective health service coverage also requires a workforce with appropriate training and skill-mix. These investments in human resources for health need to be complemented by a broader functional health system with adequate supply of medicine and consumables, functioning equipment, and effective supervision and management.^5^

## Aim

In this paper, we aim to describe and analyse trends in staff-to-baby ratios, and availability, coverage, and skill-mix of staff in 65 level-2 neonatal units in Kenya, Malawi, Nigeria, and Tanzania.

## Methods

### NEST360

The Newborn Essential Solutions and Technologies (NEST360) Alliance is working with governments in Kenya, Malawi, Nigeria, and Tanzania to reduce newborn deaths in hospitals by implementing a co-created health systems package with innovative technologies, education for clinicians and engineers, and data systems and dashboards to drive quality of care.^19–21^ The first phase of the programme, which operated from 2019-2023, has been implemented in 66 neonatal units with 13 in Kenya, 37 in Malawi, 9 in Nigeria, and 7 in Tanzania (**Figure 1**).

**Figure 1:**
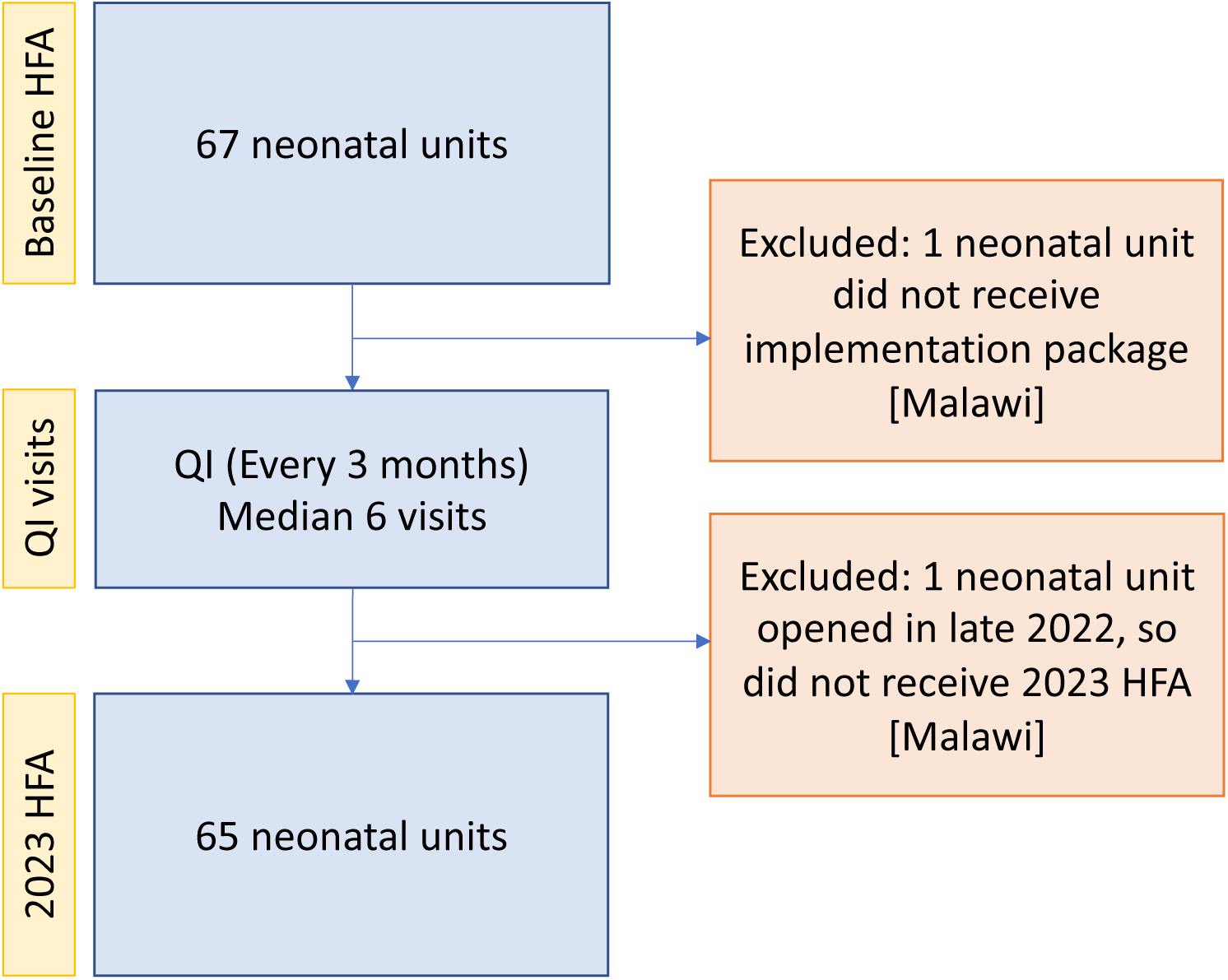
Flow figure showing number of neonatal units with Health Facility Assessment and Quality Improvement data at each time point. **Abbreviations**: HFA – Health Facility Assessment; QI – Quality Improvement

### Data Sources and Data Collection

#### Health Facility Assessment

The Health Facility Assessment (HFA) tool for SSNC, co-designed by the NEST360 Alliance and United Nations Children’s Fund (UNICEF) in partnership with four African governments, was systematically developed using a three-step evidence-based process.^22^ The tool includes items across six adapted WHO Health System Building Blocks (HSBBs) (infrastructure, medical devices and supplies, human resources, information systems, family centred care, and governance). The human resources module includes assessment of facility-level and neonatal unit staffing, facility policies and working conditions, clinical care policies, readiness to provide newborn care clinical competencies, and staff training and supervision.^23^ HFA data collection was completed in one day at each hospital using a mobile REDCap application on Android tablets.^24^ Data collectors were trained for four days before data collection began. Data were verified by HFA team supervisors and the NEST360 country database manager. All HFA data were uploaded to and stored on servers of the designated country partner during and after data collection. De-identified HFA data were transferred to a central pooled database for analysis.

### Quality Improvement Visits

The Quality Improvement (QI) visits for SSNC were led by governments with support from NEST360 to identify priorities for QI and design action plans for hospital- and ward-level improvement. QI visits took place after installation, beginning in mid-2021, and were organised every 3 months. The QI visits focused on four health systems tracks: clinical care, biomedical maintenance, information systems, and governance and leadership. Though QI visits focused on identifying areas for improvement, some process indicators were also collected, including bed occupancy and availability of neonatal unit staff by cadre. QI data collection changed over time; as such, only the first full year of QI data (2022) are included in these analyses. QI data were collected on paper and data were subsequently entered into a structured Microsoft Excel worksheet with built-in data quality checks, including validation ranges. De-identified QI data were transferred to a central pooled database for analysis.

### Context Tracker

The context tracker tool was designed to collect data on events that impact care on the neonatal unit, including loss of power or water, nurse rotations and clinical staff strikes, consumable and medicine stockouts, natural disasters, and civil unrest. Data were collected on paper weekly after installation, and data were subsequently entered into the web-based REDCap application.^24^ De-identified context tracker data were transferred to a central pooled database for analysis.

#### Data Inclusion and Indicators

Clinical staffing data were collected during each HFA and QI visit. Hospital-level and other health systems data, as well as other staff coverage data, were only collected during each HFA visit (**Table 1**). Hospital data included total number of neonatal units at each hospital (some hospitals have separate newborn wards on the same hospital campus). Detailed questions asked and response options can be found in the HFA tool on the implementation toolkit for small and sick newborn care.^21,25^

**Table 1:**
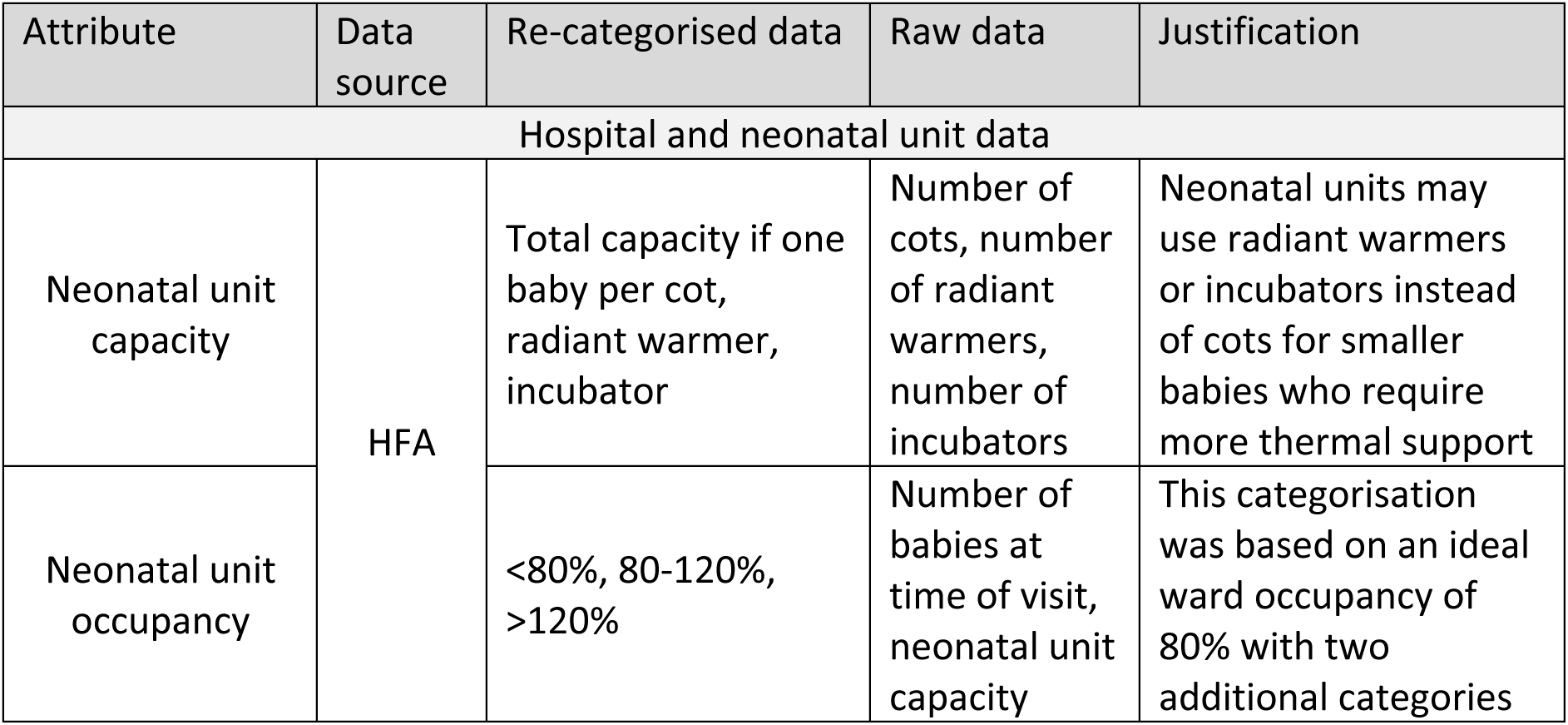
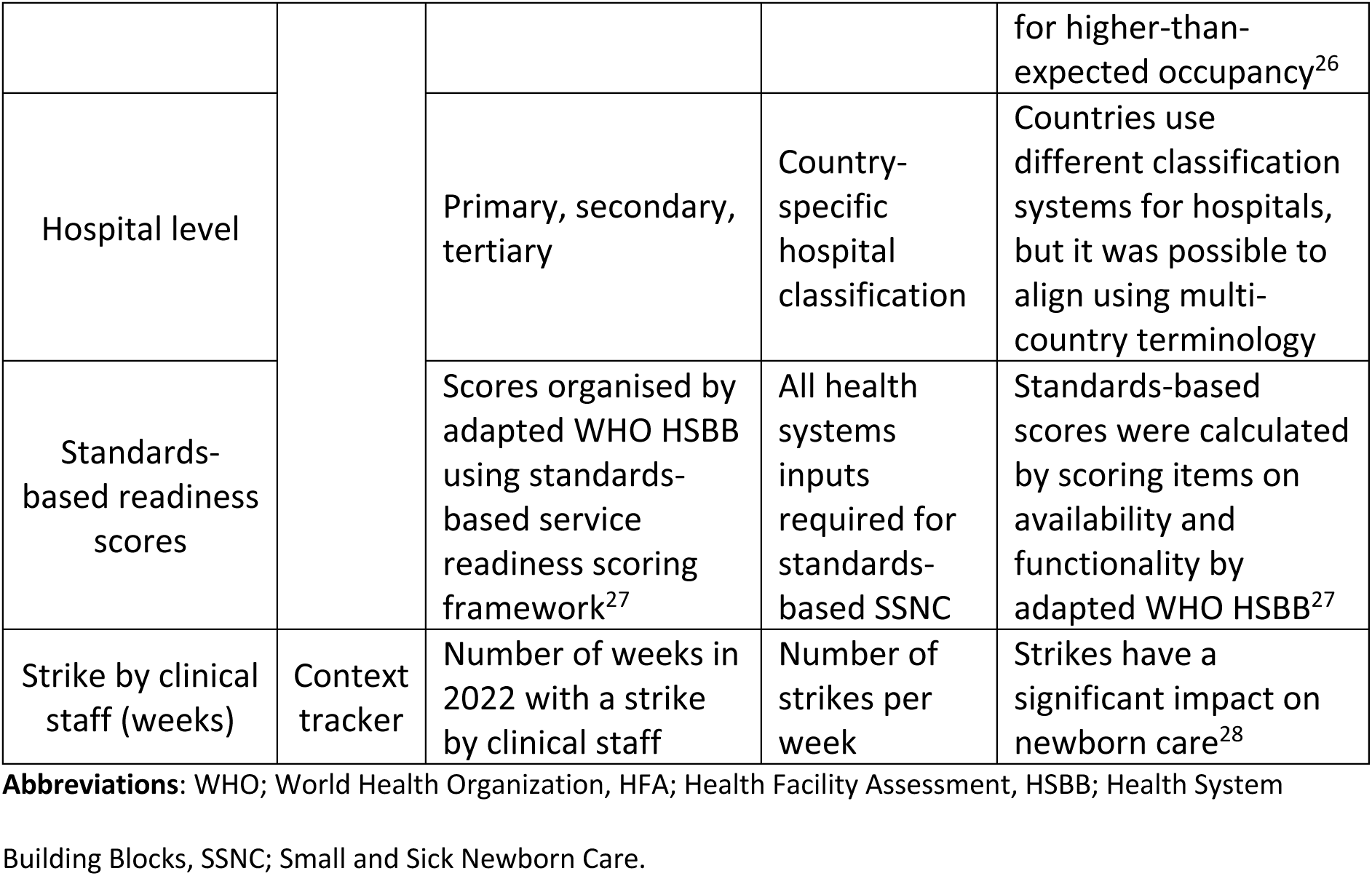
Hospital and neonatal unit data and indicators.

| Attribute | Data source | Re-categorised data | Raw data | Justification |
| --- | --- | --- | --- | --- |
| Hospital and neonatal unit data |  |  |  |  |
| Neonatal unit capacity | HFA | Total capacity if one baby per cot, radiant warmer, incubator | Number of cots, number of radiant warmers, number of incubators | Neonatal units may use radiant warmers or incubators instead of cots for smaller babies who require more thermal support |
| Neonatal unit occupancy |  | <80%, 80-120%, >120% | Number of babies at time of visit, neonatal unit capacity | This categorisation was based on an ideal ward occupancy of 80% with two additional categories for higher-than-expected occupancy<sup>26</sup> |
| Hospital level |  | Primary, secondary, tertiary | Country-specific hospital classification | Countries use different classification systems for hospitals, but it was possible to align using multi-country terminology |
| Standards-based readiness scores |  | Scores organised by adapted WHO HSBB using standards-based service readiness scoring framework<sup>27</sup> | All health systems inputs required for standards-based SSNC | Standards-based scores were calculated by scoring items on availability and functionality by adapted WHO HSBB<sup>27</sup> |
| Strike by clinical staff (weeks) | Context tracker | Number of weeks in 2022 with a strike by clinical staff | Number of strikes per week | Strikes have a significant impact on newborn care<sup>28</sup> |

**Abbreviations:** WHO; World Health Organization, HFA; Health Facility Assessment, HSBB; Health System Building Blocks, SSNC; Small and Sick Newborn Care.

#### Clinical staff data

Clinical staff categories included nurses, doctors, and clinical officers. For each staff cadre, the number of staff providing care on the day of the visit and the night before the visit was recorded. In addition, the number of staff providing ward-specific care only on the neonatal unit was recorded at each time point and for each cadre.

Nurses and doctors were further disaggregated by training and specialisation (**Table 2**). Students on clinical placement without a formal qualification were excluded from all staff counts, and counted separately for each cadre. It was not possible to disaggregate employed and locum staff on the ward the day and night before the visit. Therefore, these counts include employed and locum staff. However, for staff allocated to the neonatal unit, employed and locum staff were counted separately.

**Table 2:** Definitions for different staff cadres included in analyses.

| Cadre | Definition |
| --- | --- |
| **Nurses** |  |
| General | Any qualified nurse. |
| Specialist | Specialist nurses included those who received specialist training in paediatric or neonatal nursing through a relevant accredited training programme in line with country standards.<sup>6</sup> |
| **Doctors** |  |
| General | Any qualified general medical doctor. Includes doctors undergoing training (e.g. training to become neonatologists), but who have not yet completed specialisation training. |
| Specialist | Fully qualified paediatricians and neonatologists were included in these specialisation counts, but any doctors undergoing specialisation training were included as general medical doctors. |
| **Other staff cadres** |  |
| Biomedical technicians | Biomedical technicians included were those who received specialist technical training in line with relevant country standards. May be employed at sub-national level for multiple hospitals. |
| Engineers | Engineers included those with degree-level qualifications in any field of engineering. May be employed at sub-national level for multiple hospitals. |
| Neonatal data clerks | Neonatal data clerks included all data clerks who were assigned to the neonatal unit. |
| Laboratory technicians | Laboratory technicians included all technicians who were assigned to the central hospital laboratory. |

#### Data Analyses

For categorical variables, the number and percentage of neonatal units were analysed overall and by country. For numeric variables, the median and interquartile range (IQR) were presented overall and by country. For all coverage data, neonatal units were excluded from the denominator for that time point if the number of staff was missing.

We also included some data on characteristics of the implementing neonatal units. Strikes by clinical staff data were also tracked and analysed weekly for each neonatal unit, and aggregated by country and overall using the median and IQR.

### Objective 1: To describe clinical staff-to-baby ratios in 65 level-2 SSNC units in Kenya, Malawi, Nigeria, and Tanzania

Clinical staff-to-baby ratios for the day of each visit and night before each visit were calculated using HFA and QI data. Nurses were counted if they were providing ward-specific care on the neonatal unit (e.g. not also on the paediatric unit) and doctors were included if they were physically present at the hospital and providing care on the neonatal unit, even if not their only place of allocation.

Clinical staff-to-baby ratios were calculated for each neonatal unit, and aggregated across neonatal units, by country, hospital level (e.g. primary, secondary, tertiary), and neonatal unit occupancy. Staff-to-baby ratios were evaluated visually using boxplots and line graphs, and formally using the median and IQR to aggregate ratios across time points. For staff cadres that were not present on the day or night of the visit, excluding ratios with zero staff may be misleading and underestimate staff workload.^29^ To calculate the staff-to-baby ratios, a ratio of one staff member to the maximum number of daily admissions by country was assumed. For boxplots, the number of neonatal units with zero staff at each time point are presented in tables associated with each relevant visualisation.

### Objective 2: To describe clinical and non-clinical staff coverage and skill-mix in support of 65 level-2 SSNC units in Kenya, Malawi, Nigeria, and Tanzania

Clinical staff skill-mix and coverage were analysed across all neonatal units using baseline and 2023 HFA data by cadre. For nurses and doctors, the number of neonatal units with at least one specialist and general staff employed were analysed, including variations focusing on staff exclusively providing care on the neonatal unit (i.e. ward-specific staff), staff who were present at the time but providing care on multiple wards, and staff who were on call. In addition, the percentage of locum staff assigned to the neonatal unit was calculated. For nurses only, the percentage of specialist nurses out of all employed nurses assigned to the neonatal unit was calculated. In Malawi, the number of neonatal units with either a doctor or clinical officer assigned to the neonatal unit was also evaluated, as hospitals in Malawi employ clinical officers to provide additional clinical support on the neonatal unit.

Non-clinical staff coverage and skill-mix for biomedical technicians, engineers, data clerks, and laboratory technicians were also evaluated. For biomedical technicians and engineers, the number of hospitals with at least one employed staff member, and the number of hospitals with staff assigned exclusively to individual hospitals, were calculated. In addition, skill-mix was analysed by separately evaluating the number of biomedical technicians and engineers employed. Neonatal data clerks were evaluated by assessing the number of neonatal units with at least one neonatal data clerk assigned to the neonatal unit. For laboratory coverage, the number of hospitals with 24-hour laboratory coverage was assessed. In addition, coverage was disaggregated by day and night coverage for weekday and weekends.

### Software

HFA data were cleaned and quality checked in REDCap, while QI data were cleaned in a structured Microsoft Excel worksheet. Additional data quality checks were performed on both datasets during analysis using Stata 17 (StataCorp LLC, Texas, United States of America).^24^ Visualisations were developed using Stata 17 (StataCorp LLC, Texas, United States of America) and R software.^30,31^

### Ethical Approval

Ethical approval was received in each country from a local institutional review board (**Additional File 1)** and the London School of Hygiene & Tropical Medicine ethics committee (no. 21892). The NEST360 Alliance data sharing agreement covered data sharing between organisations. No individual consent was required for the study as no personal identification data were included.

## Results

Data from 65 neonatal units were analysed, including 36 neonatal units in Malawi, 13 in Kenya, 7 in Tanzania, and 9 in Nigeria. Data were included from two HFA and four QI time points per neonatal unit on average. 35 neonatal units were at secondary and tertiary hospitals. The remaining 30 neonatal units were at primary hospitals all of which were in Malawi (**Table 3**). Median standards-based HFA scores were 51% (IQR 48-58%) at baseline and 60% (IQR 54-66%) at 2023 HFA visits. Median time between baseline and 2023 HFAs was 31 months (IQR 29-34 months). Most neonatal units (n=44, 68%) had ideal neonatal unit occupancy of less than 80% at 2023 HFAs; however, more than half of hospitals in Kenya (n=7, 54%) were over occupied (>80%). In 2022, most hospitals had zero strike weeks; however, in Kenya, hospitals had one strike week on average in 2022.

**Table 3:** Hospital characteristics of neonatal units implementing with NEST360 at 65 neonatal units in Kenya, Malawi, Nigeria, and Tanzania.

|  | Overall | Malawi | Kenya | Tanzania | Nigeria |
| --- | --- | --- | --- | --- | --- |
| Neonatal Units | 65 | 36 | 13 | 7 | 9 |
| Baseline (2019-21) median standards-based scores [IQR] | 51%<br>[48-58%] | 49%<br>[46-51%] | 62%<br>[59-66%] | 55%<br>[53-62%] | 50%<br>[42-58%] |
| 2023 HFA median standards-based scores [IQR] | 60%<br>[54-66%] | 55%<br>[51-58%] | 66%<br>[66-67%] | 70%<br>[66-71%] | 63%<br>[50-66%] |
| Median strike weeks / weeks documented (2022) [IQR] | 0 / 44<br>[0-0] | 0 / 49<br>[0-0] | 1 / 34<br>[0-1] | 0 / 31<br>[0-0] | 0 / 40<br>[0-0] |
| Level |  |  |  |  |  |
| Tertiary/Secondary | 35 | 6 | 13 | 7 | 9 |
| Primary | 30 | 30 | 0 | 0 | 0 |
| Occupancy (number of neonatal units) |  |  |  |  |  |
| <80% | 44 | 26 | 6 | 5 | 7 |
| 80-120% | 13 | 7 | 2 | 2 | 2 |
| >120% | 8 | 3 | 5 | 0 | 0 |

**Abbreviations:** IQR; Interquartile Range, HFA; Health Facility Assessment.

### Objective 1: To describe clinical staff-to-baby ratios in 65 level-2 SSNC units in Kenya, Malawi, Nigeria, and Tanzania

#### Nurse-to-baby ratios

By 2023 HFAs, only 3 (5%) neonatal units had zero neonatal ward-specific nurses compared to 8 (12%) at baseline during the day. Across all HFA and QI time points, on average, all neonatal units had at least one nurse providing ward-specific care on the neonatal unit during the day with similar patterns during the night (**Figure 2, Additional File 2**).

**Figure 2:**
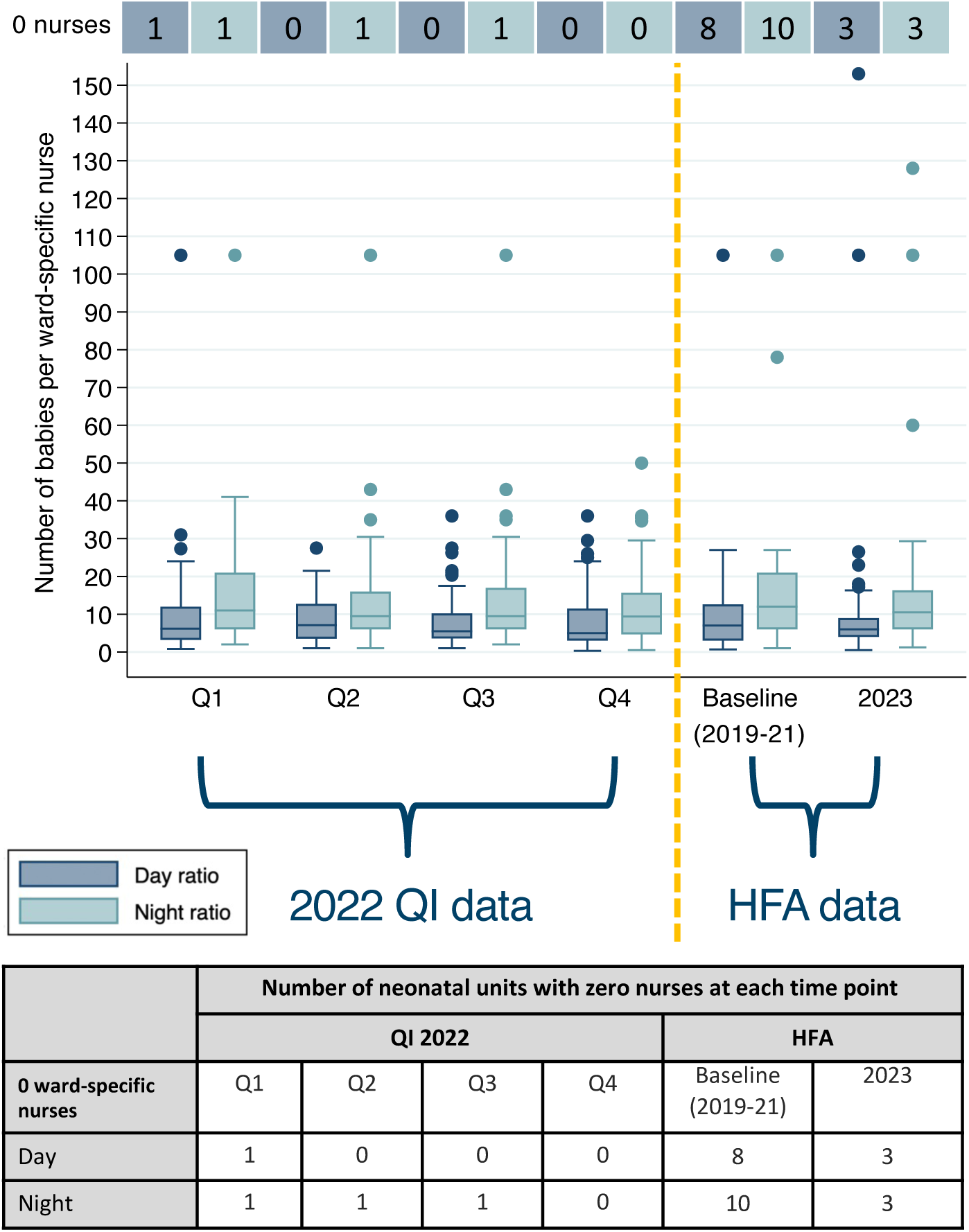
Neonatal ward-specific nurse-to-baby ratios on the day of visit and night before the visit over time at 65 neonatal units in Kenya, Malawi, Nigeria, and Tanzania. **Note:** Table displays number of neonatal units with zero ward-specific nurses at each time point. **Abbreviations:** Q; Quarter, HFA; Health Facility Assessment, QI; Quality Improvement.

Across all HFA and QI time points, median nurse-to-baby ratios were 1:6 (IQR 1:3-1:11) during the day of visit and 1:10 (IQR 1:6-1:17) the night before the visit. Nurse-to-baby ratios were worse (lower) at night compared to during the day across all time points. Median nurse-to-baby ratios varied by country, and day ratios were 1:5 (1:3-1:8) for Malawi, 1:13 (1:8-1:20) for Kenya, 1:10 (IQR 1:6-1:14) for Tanzania, and 1:3 (IQR 1:2-1:4) for Nigeria. Night ratios had similar variations by country. Though only 2022 QI data are presented given changes in data collection, ratios observed in 2022 were similar across all QI visits.

Across all hospitals, nurse-to-baby ratios during the day were worse where occupancy was greater than 120% capacity (Median 1:14, IQR 1:8-1:20) and 80-120% capacity (Median 1:10, IQR 1:7-1:14) compared to less than 80% capacity (Median 1:4, IQR 1:2-1:8) (**Figure 3, Additional File 3**). This trend was also observed for nurse-to-baby ratios during the night where even lower nurse-to-baby ratios were observed.

**Figure 3:**
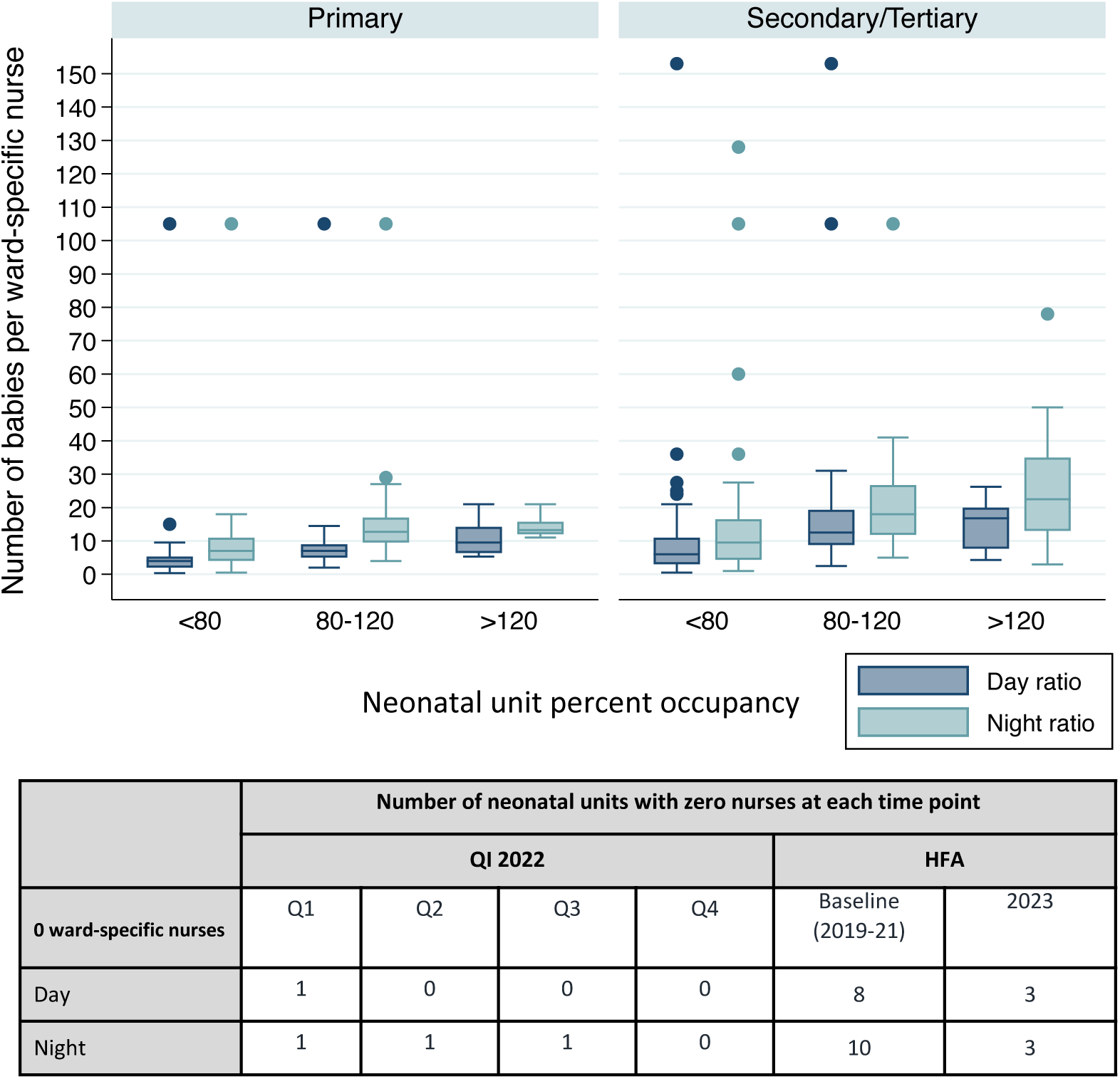
Boxplot of neonatal ward-specific nurse-to-baby ratios by hospital level (i.e. primary, secondary/tertiary) and occupancy over time at 65 neonatal units in Kenya, Malawi, Nigeria, and Tanzania. **Note:** Table displays number of neonatal units with zero ward-specific nurses at each time point. **Abbreviations:** Q; Quarter, HFA; Health Facility Assessment, QI; Quality Improvement.

Across all hospitals, nurse-to-baby ratios during the day were worse for secondary and tertiary hospitals (Median 1:9, IQR 1:4-1:14) than for primary hospitals (Median 1:4, IQR 1:3- 1:6) with similar trends at night. Some secondary and tertiary hospitals had much worse ratios, with one hospital having a nurse-to-baby ratio of 1:78 at night, likely reflecting higher admissions at these hospitals compared to primary hospitals.

#### Doctor-to-baby ratios

Across all HFA and QI time points analysed, nearly half of neonatal units (n=26, 40%) did not have a doctor who was physically present at the hospital and providing care on the neonatal unit during the day (IQR 25-29) or during the night (Median 34, IQR 33-46) on average (**Figure 4)**. When accounting for doctors who were not physically present, but were on call to provide support as needed, there were a similar number of hospitals lacking coverage by a doctor during the day and night (**Figure 5, Additional File 4**).

**Figure 4:**
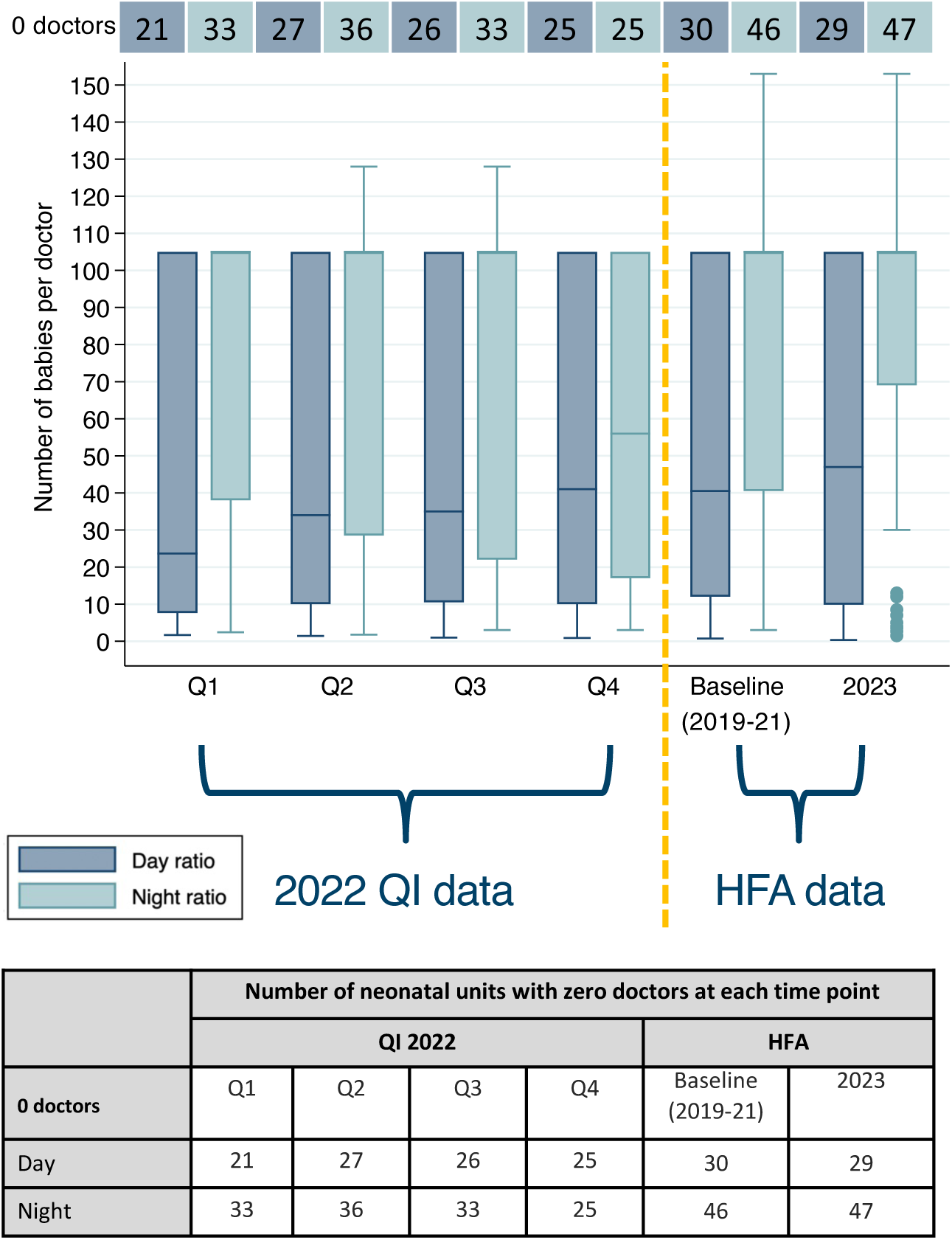
Doctor-to-baby ratios on day of visit and night before the visit over time at 65 neonatal units in Kenya, Malawi, Nigeria, and Tanzania. Abbreviations: Q; Quarter, HFA; Health Facility Assessment, QI; Quality Improvement.

**Figure 5:**
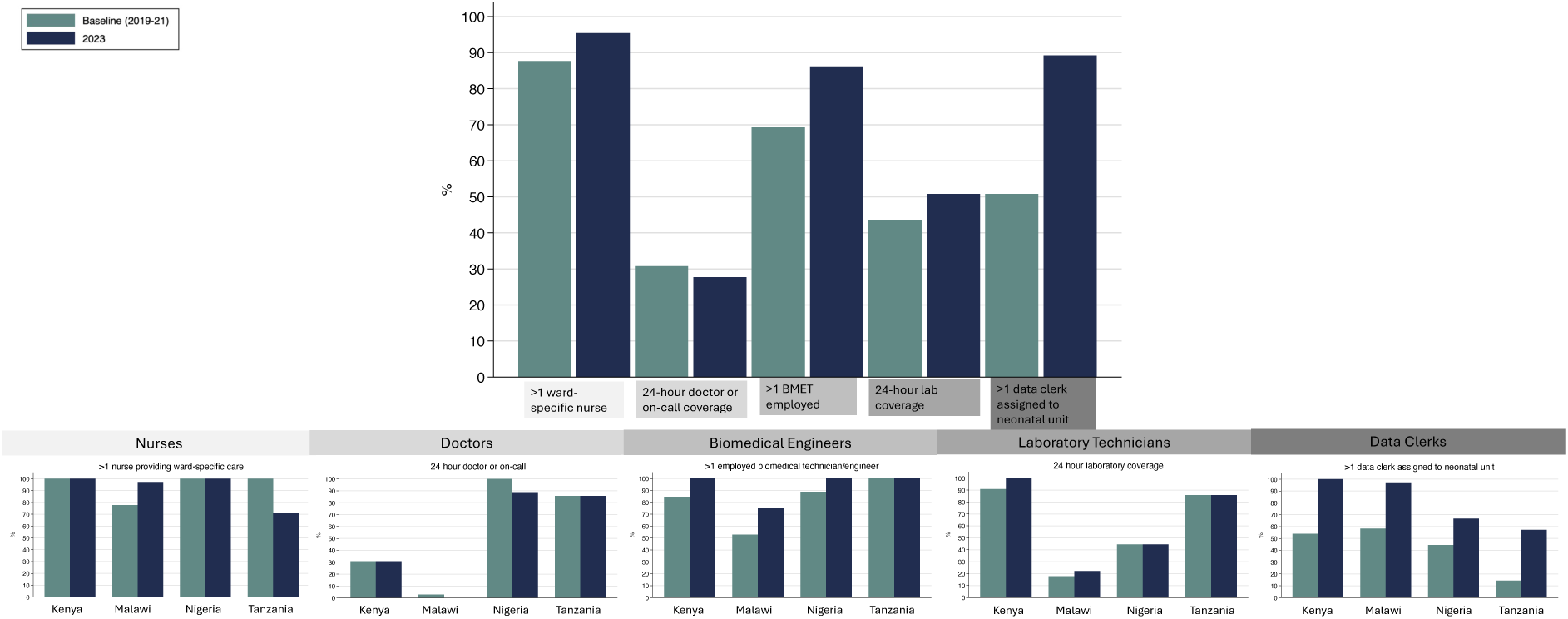
Clinical and non-clinical staff coverage and skill-mix at 65 neonatal units in Kenya, Malawi, Nigeria, and Tanzania. - Specialist doctors include paediatricians and neonatologists.
- Specialist nurses include nurses with paediatric or neonatal training as relevant for country guidelines.
- Cells show number of neonatal units (percentage).
- *Missing data for 10 neonatal units that completed different version of baseline HFA tool (n=28 for Malawi, n=11 for Kenya). Total denominator includes 55 neonatal units.
- ^±^Shows daytime staffing numbers only.
- HFA: Health Facility Assessment.
- Detailed coverage and skill-mix data can be found in Additional File 4.

Overall, median doctor-to-baby ratios were 1:35 (IQR 1:10-1:105) during the day of visit and 1:105 (IQR 1:33-1:105) the night before the visit. Median doctor-to-baby ratios varied by country, and day ratios were 1:105 (1:72-1:105) for Malawi, 1:20 (1:14-1:33) for Kenya, 1:10 (IQR 1:7-1:21) for Tanzania, and 1:2 (IQR 1:2-1:4) for Nigeria. Night ratios had similar variations by country.

### Objective 2: To describe clinical and non-clinical staff coverage and skill-mix in support of 65 level-2 SSNC units in Kenya, Malawi, Nigeria, and Tanzania

#### Nurse coverage and skill-mix

At nearly all hospitals, there was at least one nurse providing ward-specific care on the neonatal unit during the day, and there were slight increases between baseline (n=57, 88%) and 2023 HFA (n=62, 95%) (**Figure 5, Additional File 4**). However, only about one third of neonatal units had at least one specialist nurse providing dedicated care on the neonatal unit during the day of the visit, and there was no change over time. In hospitals in Kenya and Nigeria, a small proportion of all nurses providing care on the neonatal unit during the day of the visit were specialist nurses, while nurses in Malawi and Tanzania were primarily general nurses (**Additional File 4**).

#### Doctor coverage and skill-mix

Of all hospitals, approximately one third of hospitals had a doctor providing care on the neonatal unit, or on-call coverage, at all times of day or night (**Figure 5, Additional File 4**). More hospitals had doctor coverage on the neonatal unit during the day; however, this still only applied to approximately half of the hospitals. In Malawi, half of hospitals (n=19, 53%) at baseline had clinical officers providing care on the neonatal unit, but no doctors. There was no change in the doctor and clinical officer coverage between baseline and 2023 HFA.

Overall, only half of neonatal units had at least one specialist doctor (paediatrician or neonatologist) assigned, and there was no change between baseline and 2023 HFA visits. However, there was an increase in the number of neonatal units where at least one specialist doctor was providing ward-specific care on the neonatal unit, with increases particularly in hospitals in Malawi and Kenya (**Additional File 4**).

#### Biomedical technician and engineer coverage and skill-mix

Between baseline and 2023 HFA visits, the proportion of hospitals with at least one biomedical technician or engineer employed increased from 69% (n=45) to 86% (n=56) with similar increases for those exclusively assigned to the hospital (**Figure 5, Additional File 4**). These improvements in biomedical technician and engineer coverage were particularly noticeable for hospitals in Malawi, where the number of hospitals with exclusive coverage doubled from baseline (n=12) to 2023 HFA visits (n=25).

#### Data clerk coverage

There were notable improvements in the number of hospitals with at least one data clerk assigned to the neonatal unit between baseline and 2023 HFAs (**Figure 5, Additional File 4**). At baseline, approximately half of hospitals had one neonatal data clerk assigned, and at 2023 follow-up, nearly 90% of hospitals had at least one neonatal data clerk employed.

#### Laboratory technician coverage

Approximately half of hospitals had laboratory coverage twenty-four hours per day at baseline, and there was no change at 2023 HFA (**Figure 5, Additional File 4**). Limited coverage during weekday and weekend nights remained a key gap in service provision. Nearly 90% of hospitals had at least some coverage during weekday and weekend days, though there was a slight decrease in the number of hospitals with daytime coverage at the 2023 HFAs.

## Discussion

Using this large, multi-country dataset across 65 neonatal units in Kenya, Malawi, Nigeria, and Tanzania, we found that median nurse-to-baby ratios were 1:6 (IQR 1:3-1:11) during the day and 1:10 (IQR 1:6-1:17) at night. Nurse-to-baby ratios during the day were worse for secondary and tertiary hospitals (Median 1:9, IQR 1:4-1:14) than for primary hospitals (Median 1:4, IQR 1:3-1:6) with similar trends at night. Nurse-to-baby ratios were higher in Nigeria (Median 1:3, IQR 1:2-1:4) compared to ratios in other countries, which may be related to low patient volume and the high cost of care at public hospitals in Nigeria. Across all time points, patient ratios were worse at night compared to during the day. Nurse-to-baby ratios were relatively consistent across all time points analysed. Although a small number of units had zero nurses on a small number of occasions, all neonatal units had at least one nurse providing ward-specific care on the neonatal unit on average during the day with similar patterns during the night, and there were improvements in this between baseline and 2023 HFAs.

Disaggregating data further by occupancy levels and country revealed some interesting patterns. Across all hospitals, nurse-to-baby ratios were worse where there was higher neonatal unit occupancy. Lower staff ratios appeared to be more of a problem in hospitals in Kenya and Tanzania compared to Malawi and Nigeria, where more hospitals had low patient volume. However, most hospitals across the countries had staff ratios worse than national nurse-to-baby ratios recommendations from other settings, which range from 1:1 to 1:6, demonstrating the need for increased nurse staffing across all hospitals.^4^

It is not possible to provide high-quality neonatal care without adequate nurse-to-baby ratios; however there are currently no global standard ratios, though there are some national examples in India and South Africa ranging from 1:2 for intensive neonatal care to 1:6 for low-dependency newborn care, and in the United Kingdom ranging from 1:1 for intensive neonatal care to 1:4 for special newborn care.^4^ Some analyses suggest that providing high-quality Continuous Positive Airway Pressure (CPAP), a lifesaving intervention in level-2+ newborn care units, is very challenging with low staffing ratios requiring nurses to care for many patients at one time.^32^ Low staffing levels also inhibit implementation of effective, low-cost interventions, such as KMC.^33,34^ A meta-review of CPAP use in paediatric settings in LMICs suggested that staff-to-patient ratios should be no lower than 1:5, though this study examined CPAP in infants, children, and adolescents.^35^

Though staff-to-baby ratios were inadequate across most of the 65 neonatal units, there may be different reasons for this and hence different solutions. For example, hospitals with higher occupancy levels demonstrated worse staff-to-baby ratios across all countries.

Hospitals that consistently have occupancy levels above 80% may need to consider changes in their bed or cot allocation, to enable official allocation of increased human resources, as well as infrastructure changes, or restructuring of existing ward areas. Hospitals with lower occupancy levels, but still poor staff-to-baby ratios, will also require additional appropriately-skilled staff.

In terms of coverage and skill-mix, there were significant improvements over time in some cadres, which are needed to support level-2+ SSNC.^5^ For example, there were notable increases in the number of hospitals with biomedical technicians or engineers employed and assigned solely to that facility. There were also notable increases in the number of neonatal units with neonatal data clerks employed by the government. NEST360 Alliance initially supported data clerks at all 65 neonatal units, though many of the data clerks in Malawi have transitioned from NEST360-supported to government-paid data clerks between baseline and 2023 HFA visits. However, there were no or limited notable changes in nurse, doctor, and clinical officer coverage and skill-mix. Even after accounting for doctors who are on-call and not physically present at the hospital, there were no changes in the number of hospitals with doctor coverage between baseline and 2023 HFA. There were no notable changes in specialist nursing coverage, and further investment is needed to create and train a specialised cadre of neonatal nurses. While countries are developing degree-granting clinical specialisation programmes, WHO recommend interim training programmes to improve clinical skill-mix in the short term.^5^

An ethnographic study conducted in three neonatal units in Kenya revealed that some care tasks were informally delegated to parents, students, and other support staff, suggesting that parents or families may need to step-in and provide care where there is high staff workload.^36^ A recent systematic review noted that high staff workload is also associated with missed nursing care tasks.^12^ Some hospitals may rely on locum staff or students who have not completed training to fill staffing coverage gaps, which can present challenges in continuity of high-quality care and clinical team cohesion. A fully functional health system should have established positions to provide effective coverage and care. In addition, neonatal units are increasingly relying on task shifting and other reallocation of resources, particularly non-clinical tasks, to address high workload.^36^ Although task shifting may improve some aspects of care, it is crucial to ensure this is regulated and reviewed, so care is safe, and parents or families are enabled, but not stressed or overloaded.

Despite some improvements over time in the 65 neonatal units, gaps remain. Improvements include fewer neonatal units with zero nurses, and more hospitals with biomedical technicians and engineers at 2023 follow-up, including technicians providing support solely at that hospital. However, there are still several hospitals, particularly in Malawi, that do not have any technicians employed. This presents challenges in providing high-quality care especially for neonates as care is particularly dependent on devices, such as bubble CPAP and phototherapy lights, which must remain functional to use for care.^32^ In addition, large gaps remain in the number of hospitals with twenty-four hour laboratory coverage. To support management of severe conditions, such as jaundice, and appropriately treat infections, especially neonatal sepsis or meningitis, it is important to have laboratory technicians and capacity for testing at all times of day and night to support therapeutic turnaround times.^37–39^ Similarly, neonatal data clerks are important to document patient- level data, which can be used to inform quality of care, and to provide timely aggregate data for data-driven decision making across health system levels.^40^

### Strengths and Limitations

This study had strengths, notably a large dataset collected at multiple time points for staff- to-baby ratios in level-2+ neonatal units. In addition, detailed data on staff coverage and skill-mix are presented both for clinical staff and other staff cadres that support newborn care, such as biomedical technicians and engineers, data clerks, and laboratory technicians. These data include several important clinical cadres, though it was not possible to present data on pre-qualified students, who likely contribute to clinical care. There are variations in pre-qualified students by level of education, years of training, and variations within and between countries, which makes it challenging to count and define pre-qualified students. However, the data also had limitations – for example all post-qualified clinical students are included in staffing counts. In addition, it is challenging to accurately capture the number and skill-mix of staff on the neonatal unit at any given time due to staff shift patterns, allocation across multiple wards, differences in staff allocation and staff who are present on the ward, and challenges with assessing the skills and education level of every individual.

However, data are reassuringly consistent over time. It is also difficult to distinguish between locum and employed staff in most staff counts, as it was not feasible to determine whether staff providing care on the unit at various time points were locum or employed.

### Implications and next steps

More timely and routine data on human resources is foundational to measure staff-to-baby ratios over time, and staff coverage and skill-mix, and this is especially key for neonatal care.

Data are needed to inform hospital and national workforce planning, and could provide supporting evidence for developing national and global standards for staff-to-baby ratios in neonatal units, a current WHO priority. In addition, hospitals may be able to incorporate routine tracking of neonatal unit occupancy and staff-to-baby ratios to inform resource allocation, and improve patient outcomes and communication.

Governments may be able to use this data to leverage additional investment in the numbers and skills of the health workforce, making the case for return on investment. For example, a recent national investment case analysis in Tanzania showed a potential return on investment of US$8-12 for every dollar invested in scale up of neonatal care, but underlined an estimated gap of 2500 nurses nationally to reach a nurse-to-baby ratio of 1:4 with a neonatal unit in 80% of districts.^41^ Nurses are the primary care providers on newborn units, and investing in neonatal nurse specialists is likely to improve the quality of neonatal care and disability-free newborn survival, which impacts parental and family wellbeing, and may improve staff satisfaction, and reduce burnout and attrition.^7–14^

Future research would be useful to explore the relationship of staff-to-baby ratios with measurable quality of care data, and most importantly with patient outcomes. In addition, it may be useful to cost scale-up of investment in human resources for health, to estimate the need and impact of scaling up staff coverage and skill-mix with appropriate staff-to-baby ratios.

## Conclusion

Neonatal survival in hospitals requires better staff-to-baby ratios, and more staff with appropriate skills. To meet the projected shortfall in the health workforce, governments must invest in training the next generation of health workers. These investments in human resources for health need to be supported by investments in space, devices, and supplies with supportive hospital governance. Every newborn deserves a chance to survive and thrive, in every hospital and irrespective of time of day.

## Data Availability

All partners in the NEST360 alliance collaborated to create and sign data sharing and transfer agreements. The dataset from this study will be accessible upon request, pending approval from the NEST360 learning network and collaborating parties.

## List of abbreviations

CPAP: Continuous Positive Airway Pressure
HFA: Health Facility Assessment
HICs: High-Income Countries
HSBB: Health System Building Block
IQR: Interquartile Range
KMC: Kangaroo Mother Care
LMICs: Low- and Middle-Income Countries
NEST360: Newborn Essential Solutions and Technologies
QI: Quality Improvement
SSNC: Small and Sick Newborn Care
SDG: Sustainable Development Goal
UHC: Universal Health Coverage
UNICEF: United Nations Children’s Fund
WHO: World Health Organization

## Declarations

### Consent for publication

Not applicable.

### Authors’ contributions

This work was done in partnership with the NEST360 alliance, and those involved in this study are recognised for their role in data collection, management, analysis, and manuscript review. All collaborators contributed to the design of the study protocol. The objective framework and methodology for this paper were developed by REP, EOO, SC, DG and JEL. The framework and methodology underwent further refinement with inputs from co-authors. REP, EOO, DG and JEL were responsible for data curation and the formal analysis of datasets. In the analysis, important clinical insights were provided by GO, OO, RT, OT, VCE, WM, NS, GS, MC, EG, DG, and JEL. Data cleaning, analysis, and visualisations were led by REP, assisted by MOO, JT, GJ, AT, EG, JHC, RK, JC, CB, SC, JEL, DG, and EOO. Country leads implementing the Health Facility Assessment trainings, data collection, and analysis insights, were OO, OD, GO, HM, RT, JS, SKN, EZ, VOO, ER, IK, DS, JW, and GS. Quality improvement data collection and management was led by REP and GJ with support from CB, JT, and EOO. The original manuscript was initially drafted by REP with support from EOO, DG, and JEL. EMM, EZ, GO, OO, MC, OT, VCE, NS, MO, RRK and WMM are all members of the NEST360 leadership team. The manuscript underwent review and revision by all authors. All authors reviewed and gave their consent to the final version of the manuscript. The authors’ views are their own, and not necessarily from any of the institutions they represent.

## Acknowledgements

First, and most importantly, we thank the newborns and their mothers whose data are at the heart of NEST360. We also thank those involved as part of the NEST360 HFA development and all the data teams, health workers, and others involved in collecting and using the data. We also would like to thank the Ministries of Health in Kenya, Malawi, Nigeria, and Tanzania, and the hospital teams, including the clinical, biomedical engineering/technician, data, and quality improvement teams. We thank the relevant administrative staff for their assistance.

Data Collection Learning Collaborative Group (additional, not named above) Mbozu Sipalo, Millicent Alooh, Steve Adudans, Dolphine Mochache, Christina Mchoma, Joseph Bilitinyu, Pius Chalamanda, Mirriam Dzinkambani, Ruth Mhango, Fanny Stevens, Joseph Mulungu, Blessings Makhumula, Loveness Banda, Charles Banda, Brian Chumbi, Chifundo Banda, Evelyn Chimombo, Nicodemus Nyasulu, Innocent Ndau, Pilirani Kumwembe, Edna Kerubo, Nyphry Ambuso, Kevin Koech, Noel Waithaka, Calet Wakhungu, Steven Otieno, Felix Bahati, Josphine Ayaga, Jedida Obure, Nellius Nderitu, Violet Mtambo, George Mkude, Mustapha Miraji, Caroline Shayo, Camilius Nambombi, Christopher Cyrilo, Temilade Aderounmu, Akingbehin Wakeel Wale, Odeleye Victoria Yemisi, Akinola Amudalat Dupe, Samuel Awolowo, Ojelabi Oluwaseun A., John Ajiwohwodoma Ovuoraye, Balogun Adeleke Mujaid, Adedoyin Fetuga, Juilana Okanlawon, Flora Awosika, Awotayo Olasupo Michael, Omotayo Adegboyega Abiodun

## Additional Files

**Additional file 1**: *Local ethical approval for the complex evaluation of the implementation of a small and sick newborn care package with Newborn Essential Solutions and Technologies (NEST360)*.

**Additional file 2**: *Ward-specific nurse-to-baby ratios on day of and night before the visit with bed occupancy over time at 65 neonatal units in Kenya, Malawi, Nigeria, and Tanzania*.

**Abbreviations**: Q; Quarter, HFA; Health Facility Assessment, Base; Baseline.

**Additional file 3:** *Boxplot of neonatal ward-specific nurse-to-baby ratios by occupancy and by country over time at 65 neonatal units in Kenya, Malawi, Nigeria, and Tanzania*.

**Additional file 4**: *Clinical and non-clinical staff coverage and skill-mix at 65 neonatal units in Kenya, Malawi, Nigeria, and Tanzania*.

## References

1. United Nations. SDG Goal 3: Ensure healthy lives and promote well-being for all at all ages. https://sdgs.un.org/goals/goal3.

2. Joint Learning Initiative. Human Resources for Health: Overcoming the Crisis. 2004.

3. World Health Organization. Health workforce requirements for universal health coverage and the Sustainable Development Goals. (Human Resources for Health Observer, 17). Geneva: World Health Organization; 2016.

4. World Health Organization. Standards for improving the quality of care for small and sick newborns in health facilities. In. Geneva, Switzerland 2020:152.

5. World Health Organization. Human resource strategies to improve newborn care in health facilities in low- and middle-income countries. 2020.

6. World Health Organization. State of the world’s nursing 2020: investing in education, jobs and leadership. Geneva 2020.

7. Tubbs-Cooley HL, Mara CA, Carle AC, Mark BA, Pickler RH. Association of Nurse Workload With Missed Nursing Care in the Neonatal Intensive Care Unit. JAMA Pediatr. 2019;173(1):44–51.

8. Genna C, Thekkan KR, Raymakers-Janssen P, Gawronski O. Is nurse staffing associated with critical deterioration events on acute and critical care pediatric wards? A literature review. Eur J Pediatr. 2023;182(4):1755–1770.

9. Gathara D, Serem G, Murphy GAV, et al. Missed nursing care in newborn units: a cross-sectional direct observational study. BMJ Qual Saf. 2020;29(1):19–30.

10. Lasater KB, Sloane DM, McHugh MD, et al. Evaluation of hospital nurse-to-patient staffing ratios and sepsis bundles on patient outcomes. Am J Infect Control. 2021;49(7):868–873.

11. McHugh MD, Aiken LH, Sloane DM, Windsor C, Douglas C, Yates P. Effects of nurse-to-patient ratio legislation on nurse staffing and patient mortality, readmissions, and length of stay: a prospective study in a panel of hospitals. The Lancet. 2021;397(10288):1905–1913.

12. Imam A, Obiesie S, Gathara D, Aluvaala J, Maina M, English M. Missed nursing care in acute care hospital settings in low-income and middle-income countries: a systematic review. Human Resources for Health. 2023;21(1):19.

13. Shin S, Park JH, Bae SH. Nurse staffing and nurse outcomes: A systematic review and meta-analysis. Nurs Outlook. 2018;66(3):273–282.

14. Stemmer R, Bassi E, Ezra S, et al. A systematic review: Unfinished nursing care and the impact on the nurse outcomes of job satisfaction, burnout, intention-to-leave and turnover. J Adv Nurs. 2022;78(8):2290–2303.

15. Marinucci F, Majigo M, Wattleworth M, Paterniti AD, Hossain MB, Redfield R. Factors affecting job satisfaction and retention of medical laboratory professionals in seven countries of Sub-Saharan Africa. Human Resources for Health. 2013;11(1):38.

16. United Nations Children’s Fund. Toolkit for Setting Up Special Care Newborn Units, Stabilisation Units and Newborn Care Corners.

17. Limpopo Initiative for Newborn Care. Norms and standards for essential newborn care. In. Chapter 2 in essential newborn care. Polokwane 2016.

18. British Association of Perinatal Medicine. Optimal arrangements for neonatal intensive care units in the UK including guidance on their medical staffing. In. A Framework for Practice. London 2021.

19. Newborn Essential Solutions and Technologies (NEST360). NEST360 Newborn Essential Solutions and Technologies: Our Plan. In:2019.

20. Newborn Essential Solutions and Technologies (NEST360). Newborn Essential Solutions and Technologies-Education-Clinical Modules. 2020.

21. Newborn Essential Solutions and Technologies (NEST360), United Nations Children’s Fund. Implementation Toolkit: Small and Sick Newborn Care. 2021; https://newborntoolkit.org.

22. Penzias RE, Bohne C, Ngwala SK, et al. Health facility assessment of small and sick newborn care in low- and middle-income countries: systematic tool development and operationalisation with NEST360 and UNICEF. BMC Pediatr. 2024;23.

23. World Health Organization. Monitoring the Building Blocks of Health Systems : a Handbook of Indicators and their Measurement Strategies. In: 2010:110.

24. Harris PA, Taylor R, Thielke R, Payne J, Gonzalez N, Conde JG. Research electronic data capture (REDCap)—A metadata-driven methodology and workflow process for providing translational research informatics support. In. Journal of Biomedical Informatics. Vol 422009:377–381.

25. Newborn Essential Solutions and Technologies (NEST360). NEST360 UNICEF Health Facility Assessment for Small and Sick Newborn Care. 2021; https://nest360.org/project/hfa/.

26. Madsen F, Ladelund S, Linneberg A. High Levels Of Bed Occupancy Associated With Increased Inpatient And Thirty-Day Hospital Mortality In Denmark. Health Affairs. 2014;33(7):1236–1244.

27. Penzias RE, Bohne C, Gicheha E, et al. Quantifying health facility service readiness for small and sick newborn care: comparing standards-based and WHO level-2+ scoring for 64 hospitals implementing with NEST360 in Kenya, Malawi, Nigeria, and Tanzania. BMC Pediatr. 2024;23.

28. Adam MB, Muma S, Modi JA, et al. Paediatric and obstetric outcomes at a faith-based hospital during the 100-day public sector physician strike in Kenya. BMJ Glob Health. 2018;3(2):e000665.

29. de la Cruz R, Kreft J-U. Geometric mean extension for data sets with zeros. 2018.

30. Stata Statistical Software: Release 16 [computer program]. College Station, TX: StataCorp LLC; 2019.

31. RStudio: Integrated Development for R [computer program]. Boston, MA, USA: RStudio; 2024.

32. Dewez JE, Nangia S, Chellani H, White S, Mathai M, Van Den Broek N. Availability and use of continuous positive airway pressure (CPAP) for neonatal care in public health facilities in India: A cross-sectional cluster survey. In. BMJ Open. Vol 102020:1–9.

33. Tumukunde VS, Katongole J, Namukwaya S, et al. Kangaroo mother care prior to clinical stabilisation: Implementation barriers and facilitators reported by caregivers and healthcare providers in Uganda. PLOS Glob Public Health. 2024;4(7):e0002856.

34. Sivanandan S, Sankar MJ. Kangaroo mother care for preterm or low birth weight infants: A systematic review and meta-analysis. medRxiv. 2022:2022.2009.2014.22279053.

35. Sessions KL, Smith AG, Holmberg PJ, et al. Continuous positive airway pressure for children in resource-limited settings, effect on mortality and adverse events: systematic review and meta-analysis. Arch Dis Child. 2022;107(6):543–552.

36. Nzinga J, McKnight J, Jepkosgei J, English M. Exploring the space for task shifting to support nursing on neonatal wards in Kenyan public hospitals. Hum Resour Health. 2019;17(1):18.

37. Iroh Tam P-Y, Bendel CM. Diagnostics for neonatal sepsis: current approaches and future directions. Pediatric Research. 2017;82(4):574–583.

38. Mwogi T, Mercer T, Tran DNT, Tonui R, Tylleskar T, Were MC. Therapeutic turnaround times for common laboratory tests in a tertiary hospital in Kenya. PLoS One. 2020;15(4):e0230858.

39. Jacobs J, Hardy L, Semret M, et al. Diagnostic Bacteriology in District Hospitals in Sub-Saharan Africa: At the Forefront of the Containment of Antimicrobial Resistance. Front Med (Lausanne). 2019;6:205.

40. Cross JH, Bohne C, Ngwala SK, et al. Neonatal inpatient dataset for small and sick newborn care in low- and middle-income countries: systematic development and multi-country operationalisation with NEST360. BMC Pediatrics. 2023;23(2):567.

41. Kamuyu R, Tarus A, Bundala F, et al. Investment case for small and sick newborn care in Tanzania: systematic analyses. BMC Pediatr. 2023;23(Suppl 2):632.

